# Self-supervised plasma proteomic representations for prospective disease prediction across varying protein availability

**DOI:** 10.64898/2026.09.10.26362789

**Authors:** Yonghyun Nam, Thomas M. Westbrook, Jakob Woerner, Jaehyun Joo, Matthew E. Lee, Michelle McKeague, Amy E. Baxter, Shwetank, Matei Ionita, Sokratis A. Apostolidis, Allison R. Greenplate, E. John Wherry, Dokyoon Kim

## Abstract

Large-scale plasma proteomics offers opportunities to characterize disease susceptibility and improve prospective risk prediction, but transferring proteomic predictors across datasets remains challenging because measured protein sets differ across cohorts, study phases and assay configurations. Here we developed a self-supervised protein-token Transformer that maps the proteins observed in each sample to a fixed-dimensional participant representation, allowing unavailable proteins to be omitted rather than imputed. Using plasma proteomic profiles from 53,014 participants in the UK Biobank Pharma Proteomics Project, we pretrained the encoder by masked-protein reconstruction and evaluated whether disease models developed from comprehensive 2,920-protein profiles could be reused with a predefined subset of 1,460 proteins. Across 144 diseases, median AUC was 0.679 with comprehensive coverage and 0.637 when the same encoder and disease models were applied to partial-coverage representations without refitting; retraining only the disease-specific models increased median AUC to 0.673. Under partial coverage, protein-token proteomic risk scores (ProRS) exceeded coefficient-truncated LASSO ProRS by a median paired AUC difference of 0.027 and were comparable to LASSO ProRS refitted using outcome labels, with a median difference of 0.003. Performance was relatively stable across cardiovascular-kidney-metabolic diseases but more heterogeneous across autoimmune diseases, while protein-token representations improved discrimination beyond clinical covariates for 10 of 12 focused diseases under partial coverage without refitting. These results support self-supervised protein-token representations as a strategy for building proteomic prediction models that remain usable across heterogeneous measurement settings.

## 1. Introduction

Large-scale population proteomic studies have enabled systematic characterization of circulating proteins and their associations with complex diseases^1,2^. Circulating proteins reflect ongoing physiological and pathological processes and can provide prospective information beyond demographic, clinical and genetic factors^3,4^. Accordingly, multivariable plasma protein models have been developed to predict a broad range of incident diseases^3-5^.

A practical limitation of conventional proteomic risk models is their dependence on a fixed set of protein predictors^5,6^. Applying a model to a new dataset generally requires that the proteins selected during development are also measured in the target population. In practice, however, both the number and identity of measured proteins may differ across cohorts, study phases, assay panels and proteomic technologies^7-10^. A model developed from proteins (a), (b), (c) and (d), for example, cannot be applied as originally specified when only (a) and (b) are available. Reconstructing the model using the available proteins requires sufficiently large samples with labeled outcomes and produces a separate model for each measurement setting. This dependence on an identical feature space limits the portability of proteomic risk models, particularly in datasets with valuable proteomic measurements but insufficient outcome data for repeated disease-specific model development (**Fig. 1a**).

**Figure 1.**
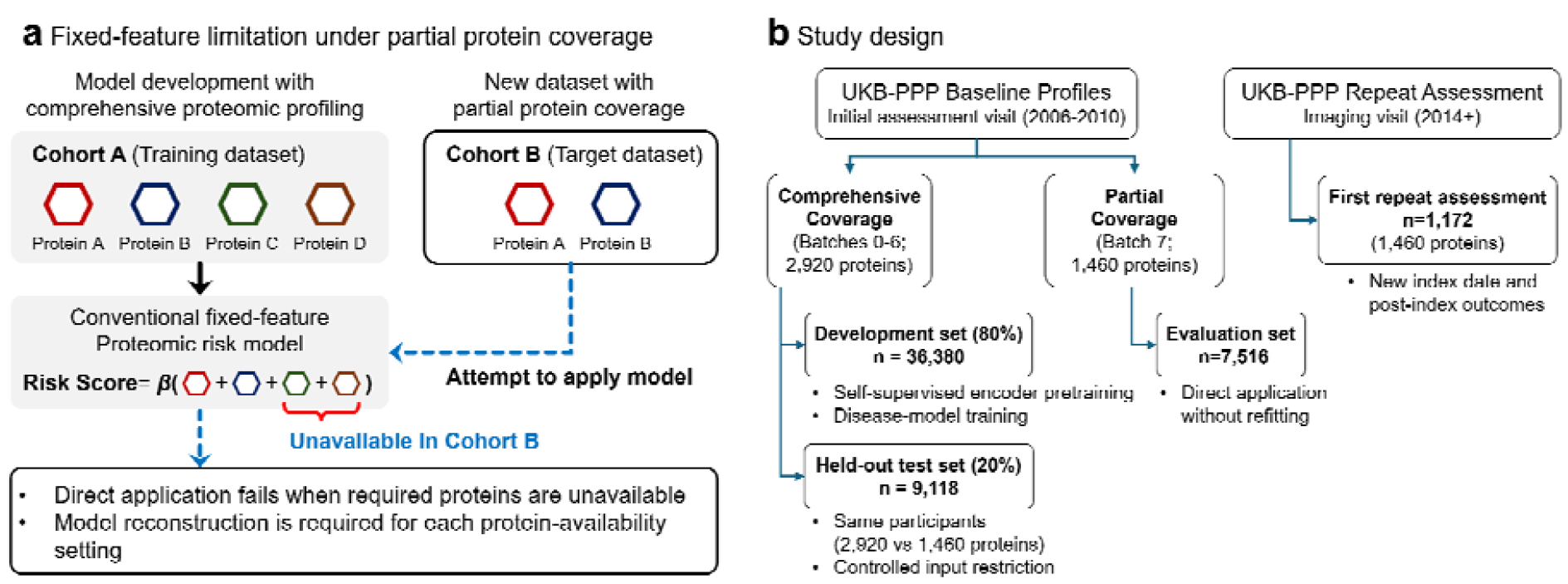
Motivation and study design. **a**, Conventional proteomic risk models depend on a fixed set of protein predictors and cannot be applied as originally specified when required proteins are unavailable. **b**, UKB-PPP comprehensive- and partial-coverage datasets used for model development, controlled input restriction, direct evaluation without refitting, and application at a later repeat assessment.

Representation learning offers a potential alternative. Rather than representing each participant as a fixed vector of protein measurements, a model can integrate the proteins observed in each sample into a common fixed-dimensional representation. Treating proteins as identifiable tokens paired with their measured abundances allows the model to process the proteins available in a given profile while omitting those without available measurements, an approach that parallels token-based self-supervised models developed for other high-dimensional molecular data^11-13^. Self-supervised learning can further capture shared structure within the plasma proteome without disease labels, enabling a single representation to support multiple downstream prediction tasks^14-16^.

Whether such representations remain useful when protein coverage changes is not established. Representations generated from partial profiles may differ from those derived from comprehensive measurements, particularly when omitted proteins carry disease-relevant information. Consequently, disease models trained on comprehensive representations may not remain valid when applied to representations generated from a smaller protein set. Addressing this question requires a controlled design in which protein coverage varies while population, phenotype definitions and follow-up are otherwise held as consistent as possible.

Here we used the UK Biobank Pharma Proteomics Project (UKB-PPP) to evaluate self-supervised plasma proteomic representation learning for prospective disease prediction under heterogeneous protein coverage. We developed a protein-token encoder using masked-protein reconstruction and used the resulting participant representations to train disease-specific proteomic risk models. Within the same biobank resource, we compared comprehensive profiles containing 2,920 proteins with profiles containing a predefined 1,460-protein subset. We evaluated whether disease models developed from comprehensive profiles could be applied without refitting after controlled restriction of protein inputs, in a separate cohort originally measured for partial coverage and in newly collected profiles from later assessments (**Fig. 1b**).

Our objective was not to establish invariance across arbitrary protein subsets, assay platforms or proteomic technologies. Rather, we asked whether self-supervised protein representations could reduce the dependence of disease-prediction models on the exact protein feature set available during development. This framework provides a step toward proteomic risk models that can operate across heterogeneous measurement settings without requiring disease-specific reconstruction whenever protein coverage changes.

## 2. Methods

To evaluate prospective disease prediction under heterogeneous protein availability, we developed a self-supervised protein-token encoder that maps variable sets of observed proteins to fixed-dimensional participant representations. The frozen encoder and disease-specific prediction models developed from comprehensive baseline profiles were evaluated after controlled restriction of protein inputs, in a separate baseline cohort with partial protein coverage, and using newly collected partial-coverage profiles at later assessments. Analyses covered 144 incident diseases, with focused evaluation of six cardiovascular-kidney-metabolic (CKM) and six autoimmune diseases (AiD) (**Fig. 1b**).

### 2.1. Study design, plasma proteomic data, and incident outcomes

We analyzed baseline plasma proteomic data from 53,014 participants in the UKB-PPP^1,8^. All baseline samples were profiled using assays from the Olink Explore 3072 platform, in which protein targets are organized into eight predefined assay panels. Batches 0–6 included all eight panels and provided measurements for 2,920 proteins after quality control, whereas batch 7 included four panels and provided measurements for 1,460 proteins. The 1,460 proteins measured in batch 7 formed a predefined subset of those measured in batches 0–6. We used this nested difference in protein coverage to define a comprehensive-coverage cohort for model development and held-out testing and a separate partial-coverage cohort for evaluation. The same 1,460-protein subset was measured at a later repeat assessment and was used to evaluate prediction from newly collected partial-coverage profiles.

We used the Normalized Protein eXpression (NPX) values provided by UKB-PPP and aligned all profiles to the common 2,920-protein manifest. Proteins without available NPX values, whether unassayed by design or missing, were omitted from the model input rather than imputed. No additional batch- or assessment-specific harmonization was performed.

The development and held-out test partitions were defined once at the participant level before model training. No held-out participant contributed any measurement to self-supervised representation learning or disease-head training. Participants and proteomic measurements were organized into four analysis datasets:

1. **Comprehensive-coverage development cohort:** 80% of participants from batches 0–6 (n=36,380). Their baseline profiles were used for self-supervised representation learning and endpoint-specific disease-head training
2. **Comprehensive-coverage test cohort:** the remaining 20% of participants from batches 0– 6 (n=9,118). These participants were held out from all model training and used for controlled comparisons in which each baseline profile was encoded using either all 2,920 proteins or the predefined 1,460-protein subset
3. **Baseline partial-coverage evaluation cohort:** participants from batch 7 (n=7,516). These participants were excluded from all model development and used to evaluate application to profiles originally measured for only the 1,460-protein subset.
4. **Repeat-assessment datasets:** partial-coverage profiles collected at the first repeat assessment (n=1,172). These profiles were not used for model training. The assessment was treated as a new prediction dataset, with the proteomic assessment date defining a new index date and post-index disease outcomes reconstructed separately.

Protein means and standard deviations were estimated in the comprehensive-coverage development cohort and applied unchanged to all other datasets.

Incident outcomes were derived using FinnGen endpoint definitions applied to linked UK Biobank health records^6,17^. The date of the proteomic assessment used for prediction served as the index date, and follow-up was censored at the earlier of death or October 31, 2022. For each endpoint, participants with a qualifying diagnosis on or before the index date were excluded from the endpoint-specific risk set.

The primary 144-endpoint analysis used a three-year prediction horizon. Incident cases were defined by a first qualifying diagnosis within three years after the index date. Controls were required to remain free of the endpoint through three years and to have at least three years of uncensored follow-up; event-free participants censored before three years were excluded. Candidate endpoints were required to have at least 100 incident cases in the comprehensive-coverage development cohort, and endpoint-level performance was reported when at least 10 incident cases were available in the comprehensive-coverage test cohort, yielding 144 evaluable endpoints. These endpoints represented a broad spectrum of incident diseases across major clinical domains, including circulatory, metabolic, musculoskeletal, digestive, respiratory, neurological, immune-mediated, neoplastic, genitourinary, and dermatological conditions.

Focused analyses examined six CKM endpoints—atrial fibrillation (AF), coronary heart disease (CHD), myocardial infarction (MI), venous thromboembolism (VTE), chronic kidney disease (CKD), and type 2 diabetes (T2D)—and six AiD endpoints—multiple sclerosis (MS), coeliac disease (CED), ulcerative colitis (UC), psoriasis (PSO), rheumatoid arthritis (RA), and systemic lupus erythematosus (SLE). To increase incident event counts for the autoimmune diseases, these analyses used all available follow-up from the baseline assessment rather than a fixed three-year horizon. Incident cases had a first qualifying diagnosis after baseline and before censoring, whereas controls remained free of the endpoint through their censoring date. Because the focused analyses used a different outcome window, their results were evaluated separately from those of the three-year 144-endpoint analysis. The same full-follow-up outcome definition was reconstructed at the first repeat assessment, using the corresponding proteomic assessment date as the new index date. Participants with the disease on or before that date were excluded, incident cases were defined by a first qualifying diagnosis after the index date and before censoring, and controls remained disease-free through censoring. Participants without observable post-index follow-up were excluded.

### 2.2. Learning plasma proteomic representations

We developed a self-supervised protein-token Transformer that maps a variable set of observed proteins to a fixed-dimensional participant representation. Let *P*= {1, …, *P*}, where *P* = 2920, denote the canonical protein vocabulary and let *O*_*i* ⊆_ *P* denote the proteins observed for participant *i*. For each observed protein *p* ∈ *O*_*i*_, the input token was defined as 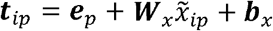 where *e*_*p*_ ∈ ℝ^256^ is a learned protein-identity embedding, 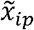 is the standardized NPX value, and 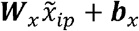 is its linear abundance projection. Proteins without available NPX values were omitted from the input sequence rather than imputed. A classification token was prepended to the observed protein tokens, and the sequence was processed by a Transformer encoder *f*_0_:

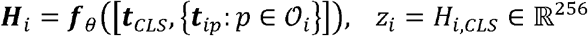

The classification-token output z_i_ was used as the participant-level plasma proteomic representation. The encoder comprised six Transformer layers, eight attention heads, a hidden dimension of 256, and a feed-forward dimension of 1,024. Because the representation dimension was fixed while the number and identity of input proteins could vary, the same encoder could process profiles with different protein coverage.

The encoder was pretrained exclusively in the comprehensive-coverage development cohort and without using disease labels^14,15^. Pretraining used masked-protein reconstruction with variation in the proteins made available to the model. For each minibatch, an availability view *V* was sampled from the complete 2,920-protein set, a random subset of 800-2,500 proteins, or the predefined 1,460-protein subset, with probabilities of 0.3, 0.4, and 0.3, respectively. The predefined 1,460-protein view was included to expose the encoder to the prespecified partial-coverage configuration evaluated in this study, whereas randomly sampled views exposed the encoder to broader variation in protein availability. The proteins supplied for participant *i* were therefore 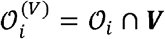.

Within each availability view, 15% of the available proteins were selected as the masked set 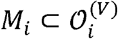. For these proteins only, the abundance projection was replaced with a learned mask embedding while the protein-identity embedding was retained. Proteins excluded from the availability view, including proteins not assayed in partial-coverage profiles, were omitted from the input sequence and were not assigned mask embeddings. A reconstruction head *g*_*ϕ*_ predicted the standardized NPX value from the corresponding Transformer hidden state. The model was optimized by minimizing mean squared reconstruction error over masked proteins:

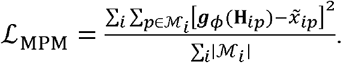

Pretraining was performed for 30 epochs using AdamW with an initial learning rate of 10^−3^, weight decay of 10^−2^, cosine learning-rate decay, and gradient clipping at 1.0. An effective batch size of 64 was obtained using a micro-batch size of eight and eight gradient-accumulation steps. Mixed-precision training was used where supported, and the encoder parameters from the final training epoch were used for downstream analysis.

After pretraining, all encoder parameters were frozen, and no disease-specific fine-tuning was performed. Representations were extracted deterministically, without masking or random protein subsampling. For the comprehensive-coverage development and test cohorts, representations were generated using either all 2,920 proteins or the predefined 1,460-protein subset, as required by the downstream analysis. For the baseline partial-coverage evaluation cohort and the repeat-assessment dataset, representations were generated from the 1,460 proteins actually measured in those profiles. The same pretrained encoder parameters were used in every setting.

### 2.3. Prospective disease prediction from frozen proteomic representations

The self-supervised encoder generated disease-agnostic participant representations but did not itself estimate disease risk. We therefore trained a separate L2-regularized logistic regression model for each disease while keeping the encoder frozen. We use proteomic risk score (ProRS) as a general term for disease-risk models incorporating plasma proteomic information^6^. The representation-based approach is referred to as protein-token ProRS, whereas the conventional model fitted directly to protein measurements is referred to as LASSO ProRS.

Protein-token ProRS combined the 256-dimensional proteomic representation with age, sex, body mass index, smoking status, and ethnicity. Models for the 144-disease analysis were trained using a common three-year prediction horizon to enable consistent comparison across diseases. Focused CKM and AiD models were trained separately using all available follow-up to increase incident event counts, particularly for autoimmune diseases, and were therefore evaluated as a separate analysis. Age and body mass index were standardized using development-cohort statistics. Missing continuous clinical covariates were mean-imputed with missingness indicators, and categorical covariates were encoded using development-defined categories. This imputation applied only to clinical covariates; proteins without available NPX values were omitted from the encoder input rather than imputed. All transformations were applied unchanged to the evaluation datasets. For the repeat-assessment analysis, separate disease-specific proteome-only models were trained using representations generated from all 2,920 proteins in the comprehensive-coverage development cohort and full-follow-up outcomes. These models allowed prediction from newly collected proteomic profiles without requiring updated clinical covariates.

We evaluated three protein-token ProRS settings:

- **Comprehensive coverage**: the disease model was trained using representations generated from all 2,920 proteins in the development cohort and evaluated in the held-out test cohort.
- **Partial coverage, no refit**: the same frozen encoder and disease model were applied to representations generated from the predefined 1,460-protein subset, without retraining, feature selection, or recalibration.
- **Partial coverage, refit**: the encoder remained frozen, but the disease model was retrained using 1,460-protein representations from the development cohort.

To benchmark against conventional fixed-feature prediction, we constructed disease-specific L1-regularized logistic regression models directly from standardized protein measurements, using the same participants, outcomes, and clinical covariates. We evaluated a comprehensive-coverage model using all 2,920 proteins, a truncated model in which coefficients for unavailable proteins were set to zero without refitting, and a partial-coverage model refitted using the 1,460-protein subset. The truncated model represented direct application of an existing fixed-feature model, whereas the refitted model represented conventional reconstruction when outcome labels were available.

Representation-based models used L2 regularization with C=1.0, and sparse models used L1 regularization with C=0.1. All models used class-balanced weighting. L2 and L1 models were fitted using the lbfgs and liblinear solvers, respectively, with maximum iteration counts of 5,000 and 1,000. Model settings were fixed across diseases and were not selected using held-out data.

To evaluate incremental predictive value beyond clinical covariates, we additionally trained disease-specific covariate-only logistic regression models using age, sex, body mass index, smoking status, and ethnicity. These models used the same development participants, outcome definitions, covariate preprocessing, regularization, and class weighting as the corresponding protein-token ProRS models. The covariate-only models were applied unchanged to the comprehensive-coverage test cohort and the baseline partial-coverage evaluation cohort. Comparisons with protein-token ProRS were paired within each evaluation dataset because predictions were generated for the same participants

### 2.4. Evaluation under partial protein coverage and at repeat assessments

The primary evaluation used the comprehensive-coverage test cohort, in which each participant was encoded using either all 2,920 proteins or the predefined 1,460-protein subset. The same comprehensive-coverage-trained disease model was applied to both representations, holding participants, outcomes, covariates, preprocessing, and model parameters constant while varying only protein availability. The frozen encoder and disease models were then applied unchanged to the baseline partial-coverage evaluation cohort, whose participants were excluded from model development. Comparisons between the two cohorts were unpaired because they comprised different participants.

The first repeat assessment was treated as a new prediction dataset. The assessment date served as the new index date, and prevalent disease exclusions and incident outcomes were reconstructed relative to that date using all available post-index follow-up. The frozen encoder and full-follow-up proteome-only disease models were applied unchanged to the measured 1,460-protein profiles without protein imputation, assessment-specific normalization, refitting, or recalibration. Disease-specific AUC was reported when at least 10 incident cases and 10 controls were available; estimates based on fewer than 30 incident cases were considered exploratory. Because some repeat-assessment participants overlapped with the baseline development cohort, this analysis was interpreted as application to newly collected proteomic profiles rather than fully independent participant-level validation.

### 2.5. Statistical analysis

Prediction discrimination was evaluated using the area under the receiver operating characteristic curve (AUC). Model comparisons within an evaluation cohort were paired because predictions were generated for the same participants. Comparisons of performance estimates between the comprehensive-coverage test cohort and the separate baseline partial-coverage evaluation cohort were unpaired because the cohorts comprised different participants. For the 144-disease analysis, disease-specific 95% confidence intervals were estimated using 500 participant-level bootstrap replicates, and confidence intervals for median AUC and median paired AUC differences across diseases were estimated by resampling diseases 2,000 times. For the focused CKM and AiD analyses, including comparisons with covariate-only models, disease-specific paired confidence intervals were estimated using 1,000 participant-level bootstrap replicates. Repeat-assessment AUC confidence intervals were estimated using 1,000 participant-level bootstrap replicates. Cross-disease summaries were interpreted descriptively because diseases shared participants.

## 3. Results

### 3.1. Prediction performance under partial protein coverage

Before examining disease-specific variation, we assessed whether disease models trained on representations generated from 2,920 proteins could be applied when only 1,460 proteins were available. Across 144 diseases evaluated for three-year incidence, the protein-token model achieved a median AUC of 0.679 with 2,920 proteins and 0.637 when the same frozen encoder and disease models were applied without refitting to 1,460-protein representations. Refitting only the disease models using 1,460-protein representations increased the median AUC to 0.673. The corresponding LASSO ProRS values were 0.654, 0.617, and 0.630, respectively (**Table 1**). Notably, protein-token ProRS also achieved a higher median AUC than LASSO ProRS under comprehensive coverage (0.679 versus 0.654), indicating that the flexibility of the learned representation did not entail an apparent loss of overall predictive discrimination.

**Table 1.**
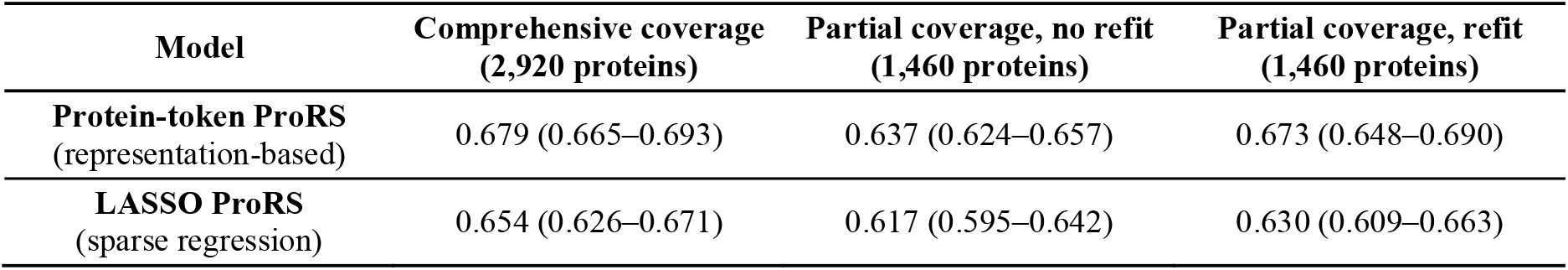
Median AUC across 144 diseases according to model and protein coverage. Values are median AUCs with descriptive 95% confidence intervals across diseases. For protein-token ProRS, refit denotes retraining the disease-specific logistic model while keeping the encoder frozen. For LASSO ProRS, no refit denotes setting coefficients for unavailable proteins to zero while retaining the original intercept and coefficients for available proteins.

The no-refit protein-token model outperformed coefficient-truncated LASSO ProRS by a median paired AUC difference of 0.027 (95% CI, 0.018–0.041) and differed from refitted LASSO ProRS by 0.003 (−0.008 to 0.013). When applied unchanged to the separate baseline cohort originally measured for 1,460 proteins, the protein-token model achieved a median AUC of 0.672, compared with 0.678 for the same diseases in the 2,920-protein test cohort. Although this comparison was unpaired and may reflect differences in batch composition or participant characteristics, it demonstrated direct application to profiles originally measured with partial protein coverage without outcome-based model refitting, recalibration, or cohort-specific normalization.

### 3.2. Disease-specific performance in CKM and autoimmune diseases

To determine whether the overall results masked disease-specific variation, we examined six CKM and six AiD diseases using all available follow-up. Incident case and control counts in the comprehensive-coverage test cohort and the baseline partial-coverage evaluation cohort are provided in **Table 2**. Among CKM diseases, applying protein-token ProRS without refitting to 1,460-protein representations reduced AUC by 0.009–0.029 relative to the 2,920-protein model (**Fig. 2a**). No-refit protein-token ProRS had a higher AUC than coefficient-truncated LASSO ProRS for all six CKM diseases, with 95% confidence intervals excluding zero for five (**Fig. 2b**). Compared with refitted LASSO ProRS, the confidence interval included zero for five diseases; type 2 diabetes was the exception, favoring refitted LASSO ProRS by 0.024 (95% CI, 0.013– 0.034; **Fig. 2c**).

**Table 2.** Incident case and control counts for the focused disease analysis. Counts represent incident cases and disease-specific controls over all available follow-up. Participants with the corresponding disease at baseline were excluded.

| CKM disease | Case / Control |  | AiD disease | Case / Control |  |
| --- | --- | --- | --- | --- | --- |
|  | Comprehensive coverage | Partial coverage |  | Comprehensive coverage | Partial coverage |
| MI | 266 / 7,907 | 225 / 6,364 | PSO | 92 / 8,972 | 72 / 7,395 |
| AF | 695 / 4,911 | 670 / 3,935 | UC | 58 / 9,018 | 48 / 7,427 |
| CHD | 328 / 8,660 | 264 / 7,071 | RA | 166 / 6,177 | 136 / 5,106 |
| CKD | 534 / 8,165 | 804 / 6,360 | CED | 40 / 8,508 | 44 / 6,972 |
| T2D | 674 / 8,204 | 517 / 6,730 | SLE | 55 / 7,519 | 18 / 6,072 |
| VTE | 277 / 8,764 | 250 / 7,204 | MS | 33 / 9,031 | 27 / 7,448 |

**Figure 2.**
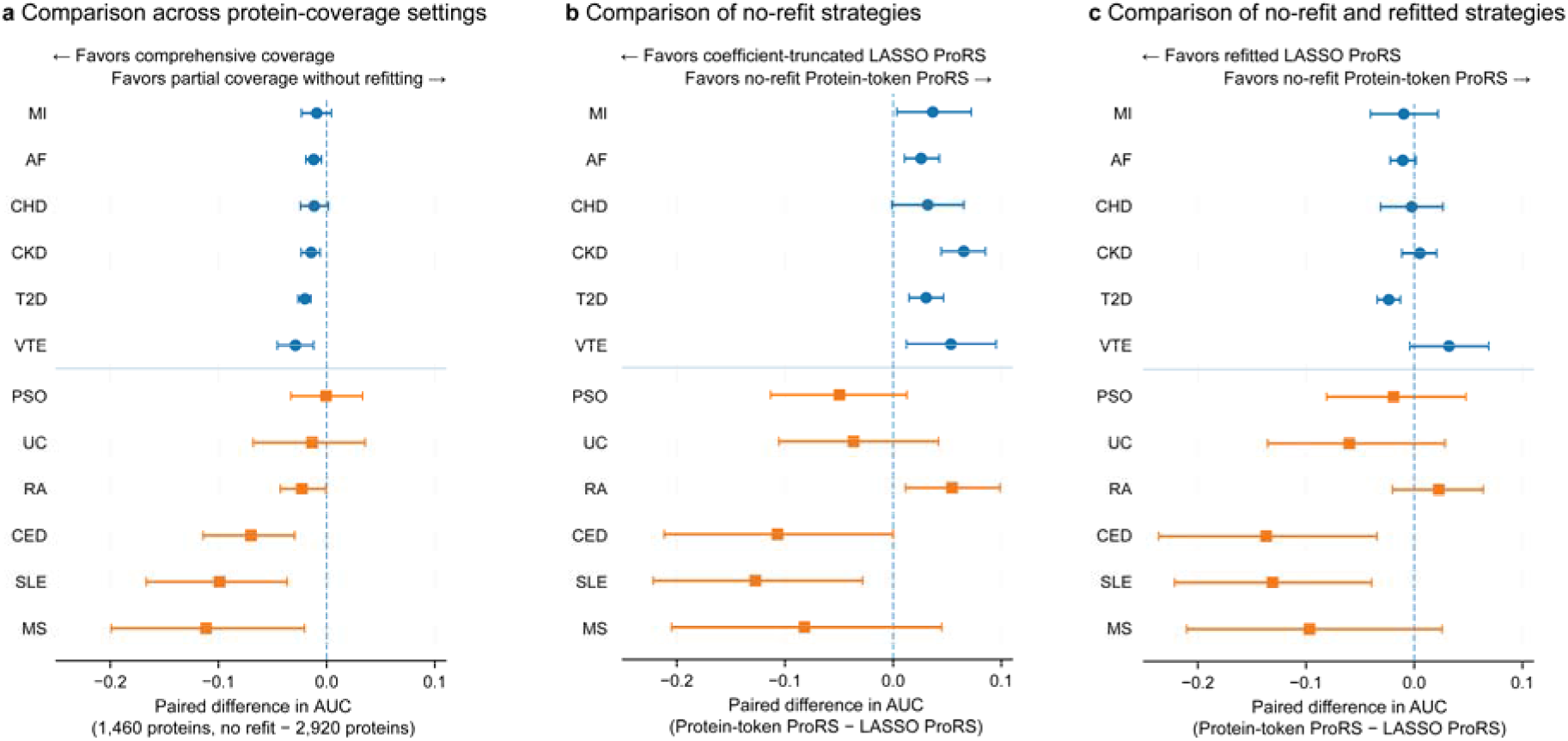
Disease-specific prediction performance under partial protein coverage. Paired AUC differences compare no-refit protein-token ProRS using 1,460 proteins with (a) protein-token ProRS using 2,920 proteins, (b) coefficient-truncated LASSO ProRS, and (c) LASSO ProRS refitted using 1,460 proteins. Points show AUC differences and horizontal lines indicate 95% bootstrap confidence intervals; positive values favor the model shown to the right.

Performance was more heterogeneous among AiD. Rheumatoid arthritis, psoriasis, and ulcerative colitis showed AUC reductions of 0.023 or less under partial coverage, whereas coeliac disease, systemic lupus erythematosus, and multiple sclerosis showed reductions of 0.070, 0.099, and 0.111, respectively, with all three confidence intervals excluding zero (**Fig. 2a**). Compared with coefficient-truncated LASSO ProRS, no-refit protein-token ProRS performed better for rheumatoid arthritis but worse for coeliac disease and systemic lupus erythematosus; confidence intervals included zero for the remaining diseases (**Fig. 2b**). Refitted LASSO ProRS also outperformed no-refit protein-token ProRS for coeliac disease and systemic lupus erythematosus, whereas the other comparisons were inconclusive (**Fig. 2c**).

When applied unchanged to the separate baseline partial-coverage cohort, protein-token ProRS achieved AUCs of 0.638–0.908 for CKM diseases and 0.545–0.776 for AiD. Because this cohort comprised different participants, these estimates cannot be interpreted as direct retention of performance relative to the comprehensive-coverage test cohort and may reflect differences in case mix and participant characteristics. Precision also varied across diseases according to the number of incident cases available in this cohort (**Table 2**). Interpretation of the separate partial-coverage estimate for systemic lupus erythematosus was limited by only 18 incident cases, whereas the paired test-cohort comparison used 55 cases. Accordingly, the paired analyses provided the primary evidence of disease-specific transfer, showing relatively stable performance across CKM diseases but greater heterogeneity among AiD.

### 3.3. Prediction at later proteomic assessments

We treated the first repeat assessment as a new prediction dataset, using the proteomic assessment date as the index date and all available post-index follow-up. The frozen encoder and prespecified proteome-only disease models were applied unchanged to the measured 1,460-protein profiles, generating finite scores for all 1,172 participants. Four CKM diseases had at least 10 incident cases and yielded exploratory AUCs ranging from 0.744 to 0.871 (median, 0.799), although none had at least 30 cases. These results demonstrate application of the fixed models to newly collected partial-coverage profiles, while limited event counts precluded robust validation.

### 3.4. Incremental predictive value beyond clinical covariates

To determine whether protein-token representations provided prospective information beyond standard clinical risk factors, we compared disease-specific models using age, sex, body mass index, smoking status, and ethnicity alone with protein-token ProRS models evaluated in the held-out comprehensive-coverage test cohort (**Fig. 3**). Adding the 2,920-protein representation increased AUC for all 12 focused diseases, with AUCs ranging from 0.616 to 0.873 compared with 0.519 to 0.818 for the covariate-only models. The descriptive median paired AUC improvement was 0.068, with particularly large gains for type 2 diabetes, chronic kidney disease, coeliac disease, and ulcerative colitis. When the same disease models were applied without refitting to representations generated from 1,460 proteins, AUC remained higher than that of the covariate-only model for 10 of 12 diseases, with a median paired improvement of 0.033. The principal exceptions were systemic lupus erythematosus, for which performance was similar to the covariate-only model, and multiple sclerosis, for which partial-coverage performance was lower. In the separate baseline partial-coverage cohort, the no-refit model also exceeded the covariate-only model for 8 of 12 diseases, with a median paired AUC improvement of 0.022. These findings indicate that protein-token representations contributed predictive information beyond the included clinical covariates, although retention of this information under partial coverage varied across diseases, particularly among autoimmune diseases.

**Figure 3.**
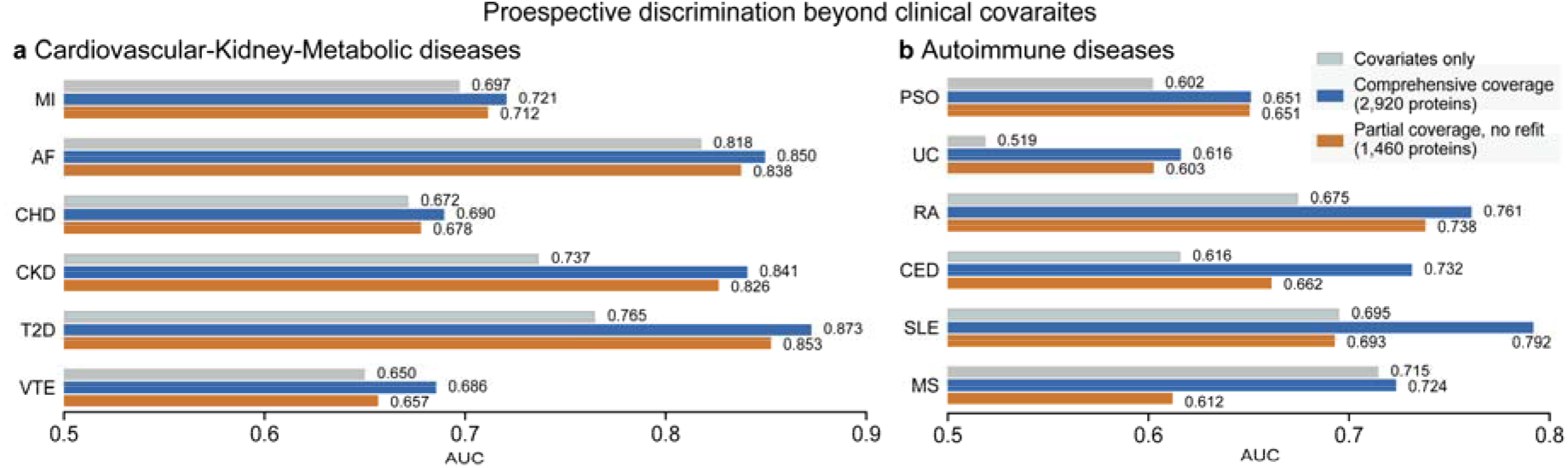
Prospective disease discrimination beyond clinical covariates. Bars show AUCs for disease-specific models containing clinical covariates alone, clinical covariates plus comprehensive-coverage protein-token representations generated from 2,920 proteins, and application of the same protein-token ProRS models without refitting to representations generated from 1,460 proteins. All models were evaluated in the held-out comprehensive-coverage test cohort using full-follow-up incident outcomes

## 4. Conclusion

We developed a self-supervised protein-token model that converts variable sets of measured plasma proteins into fixed-dimensional participant representations for prospective disease prediction. Across 144 diseases, protein-token ProRS retained useful discrimination when applied without refitting to representations generated from 1,460 rather than 2,920 proteins. Under partial coverage, the representation-based approach outperformed direct coefficient truncation of conventional LASSO ProRS and approached the performance of models reconstructed using outcome labels. Under comprehensive coverage, protein-token ProRS also achieved a higher median AUC than raw-protein LASSO ProRS, suggesting that increased portability did not require a corresponding reduction in predictive performance. The fixed encoder and disease models could also be applied directly to a separate cohort originally profiled for the 1,460-protein subset and to profiles collected at a later assessment, without protein imputation, cohort-specific normalization, or outcome-based recalibration.

Portability under partial protein coverage was disease dependent. Performance was comparatively stable across CKM diseases, whereas coeliac disease, systemic lupus erythematosus, and multiple sclerosis showed greater losses when protein inputs were restricted. Protein-token representations also improved discrimination beyond age, sex, body mass index, smoking status, and ethnicity for most focused diseases. These findings indicate that a common representation space can reduce dependence on an identical input feature set, although it cannot fully recover information carried by unmeasured proteins.

This study was limited to nested panels from the same biobank and proteomic platform. Because the predefined 1,460-protein subset was included during self-supervised pretraining, the analysis evaluated panel-aware representation learning rather than zero-shot transfer to an unseen protein configuration. Comparisons with the separate partial-coverage cohort were unpaired, and limited post-assessment events precluded robust validation at the repeat assessment. We also focused on discrimination and did not evaluate calibration, decision utility, or larger platform-specific measurement shifts.

Future work should evaluate transfer to substantially smaller and previously unseen protein panels, compare omission-based tokenization with imputation and other missing-feature strategies, and test cross-platform portability across independent cohorts profiled using Olink^7^, SomaScan^9^, or Nomic nELISA^10^. Extending representation learning across multiple assay technologies may help distinguish robustness to incomplete protein coverage from robustness to platform-specific measurement shifts. Evaluation of calibration and clinical utility will also be needed before application in prospective precision-medicine settings.

Overall, these results support protein-token representation learning as a practical strategy for developing proteomic risk models that are less tightly coupled to the exact protein feature set used during training. By allowing a fixed encoder and disease model to operate on newly observed protein subsets, the framework may reduce the need to reconstruct disease-specific models whenever protein coverage changes.

## Data Availability

The data used in this study are available through the UK Biobank upon successful application and approval. This study was conducted under UK Biobank Application 32133.

## Acknowledgements

This work was supported by NIGMS R01 GM138597 and NHLBI R01 HL169458. Use of the UK Biobank Resource in the current study was approved under Application Number [32133].

## Notes

### Competing Interest Statement

The authors have declared no competing interest.

### Author Declarations

The North West Multi-centre Research Ethics Committee of the National Health Service gave ethical approval for the UK Biobank (REC reference: 11/NW/0382).This study was conducted under UK Biobank Application 32133

